# Detection of distant familial relatedness in biobanks for identification of undiagnosed carriers of a Mendelian disease variant: application to Long QT syndrome

**DOI:** 10.1101/2023.04.19.23288831

**Authors:** Megan C. Lancaster, Hung-Hsin Chen, M. Benjamin Shoemaker, Matthew R. Fleming, James T. Baker, Hannah G. Polikowsky, David C. Samuels, Chad D. Huff, Dan M. Roden, Jennifer E. Below

## Abstract

**Importance:** The diagnosis and study of rare genetic disease is often limited to referral populations, leading to underdiagnosis and a biased assessment of penetrance and phenotype.

**Objective:** To develop a generalizable method of genotype inference based on distant relatedness and to deploy this to identify undiagnosed Type 5 Long QT Syndrome (LQT5) rare variant carriers in a non-referral population.

**Participants:** We identified 9 LQT5 probands and 3 first-degree relatives referred to a single Genetic Arrhythmia clinic, each carrying D76N (p.Asp76Asn), the most common variant implicated in LQT5. The non-referral population consisted of 69,879 ancestry-matched subjects in BioVU, a large biobank that links electronic health records to dense array data. Participants were enrolled from 2007-2022. Data analysis was performed in 2022.

**Exposures:** We developed and applied a novel approach to genotype inference (Distant Relatedness for Identification and Variant Evaluation, or DRIVE) to identify shared, identical-by-descent (IBD) large chromosomal segments in array data.

**Main Outcomes and Measures:** We sought to establish genetic relatedness among the probands and to use genomic segments underlying D76N to identify other potential carriers in BioVU. We then further studied the role of D76N in LQT5 pathogenesis.

**Results:** Genetic reconstruction of pedigrees and distant relatedness detection among clinic probands using DRIVE revealed shared recent common ancestry and identified a single long shared haplotype. Interrogation of the non-referral population in BioVU identified a further 23 subjects sharing this haplotype, and sequencing confirmed D76N carrier status in 22, all previously undiagnosed with LQT5. The QTc was prolonged in D76N carriers compared to BioVU controls, with 40% penetrance of QTc ≥ 480 msec. Among D76N carriers, a QTc polygenic score was additively associated with QTc prolongation.

**Conclusions and Relevance:** Detection of IBD shared chromosomal segments around D76N enabled identification of distantly related and previously undiagnosed rare-variant carriers, demonstrated the contribution of polygenic risk to monogenic disease penetrance, and further established LQT5 as a primary arrhythmia disorder. Analysis of shared chromosomal regions spanning disease-causing mutations can identify undiagnosed cases of genetic diseases.

## Introduction

Long QT syndrome (LQTS) is a well-recognized, rare cause of syncope and sudden cardiac death (SCD) with an estimated prevalence of 1:2000.^1^ *KCNE1* mutations cause Type 5 Long QT Syndrome (LQT5),^2^ a subtype accounting for 1-2% of autosomal dominant congenital LQTS cases. *KCNE1* encodes a function-modifying subunit for the voltage-gated slow delayed rectifier potassium current I_Ks_,^3^ and possibly other potassium currents.^4-6^ Functional studies of the missense mutation, rs74315445, resulting in p.Asp76Asn or D76N, have shown a dominant negative effect to reduce I_Ks_.^2^ However, recent registry and referral center-based studies argue that *KCNE1* variants have low penetrance (10-30%) and are not truly disease-causing, but rather function-modifying, and predispose to drug-induced forms of LQTS.^7-9^

An international consortium of 26 centers identified 89 probands with possible LQT5, 140 additional carrier relatives, and 19 cases of Jervell-Lange-Nielsen Syndrome attributed to homozygous or compound heterozygous *KCNE1* loss of function variants.^10^ The commonest mutation was D76N, with 35 probands and 63 carrier relatives. Nine probands, as well as 3 carrier relatives, were identified at Vanderbilt University Medical Center (VUMC), representing a marked enrichment relative to other sites. We hypothesized that these local probands were distantly related, and that this interrelatedness would provide an opportunity to identify additional carriers in a regional biobank and to establish the impact of D76N.

Most data on the impact of rare variants in Mendelian disease genes have been gathered in referral or registry populations. This approach overestimates true population impact, which is better assessed in large non-referral population cohorts, such as biobanks. Since most biobanks recruit participants regionally, there is often significant undocumented (“cryptic”) relatedness among participants. This oversampling of related individuals provides an abundance of genomic segments that are shared without recombination due to common ancestry. These identical-by-descent (IBD) segments provide an opportunity to study ungenotyped or poorly genotyped rare variants harbored within them. Because rare variants within IBD segments are shared if present in the common ancestor, IBD segments can identify likely carriers of rare variants,^11^ as well as inform relationships and reconstruct pedigrees.^12-15^

To generate unbiased estimates of the role of D76N and identify likely carriers in BioVU, we developed a novel approach, DRIVE (Distant Relatedness for Identification and Variant Evaluation) that leverages IBD. First, we estimated the genome-wide relatedness among the 12 clinical D76N carriers and reconstructed pedigrees. We then identified the shared haplotypes spanning *KCNE1*. We used DRIVE to identify BioVU subjects who share IBD segments containing *KCNE1* with clinic carriers, and confirmed D76N carrier status via sequencing. Finally, we assessed ECGs and medical records for features of LQTS. This enlarged carrier group in a hospital-based population improved power to revisit the debate about the role of *KCNE1* in LQTS, as well as to identify the interaction between D76N carriage and a QTc polygenic risk score (PRS) in modifying D76N penetrance. Our findings highlight the utility of IBD analysis in large biobanks to identify undetected carriers of rare pathogenic variants. This enables analyses of interacting effects, expansion of clinical phenotypes, and estimation of variant penetrance in a non-referral clinical population.

## Methods

Nine probands and three family members from the Genetic Arrhythmia Clinic at VUMC were clinically genotyped and found to be heterozygous carriers of the *KCNE1* D76N variant. Clinic samples were genotyped on MEGA^EX^ to generate haplotype data using the same array and following the same protocols as those in BioVU. Written consent was obtained under VUMC IRB approval. BioVU is the VUMC biorepository linking deidentified electronic health records (EHRs) to over 300,000 DNA samples derived from specimens about to be discarded after clinical testing.^16^ Currently in BioVU, 95,124 individuals have been genotyped on the Illumina Expanded Multi-Ethnic Genomic Array (MEGA^EX^), and 54,347 of these have had at least one ECG recorded. (See eMethods for details on genotype quality control (QC) in clinic and BioVU subjects.) SHAPEIT4^17^ was used to phase in genotype data from both BioVU EA subjects and clinic samples, separately. Genome-wide IBD proportions were calculated using the method-of-moments estimation function in PLINK^18^ after removing ancestry-informative SNPs in PRIMUS.^14^ The length and distribution of IBD segments genome-wide were used to identify more distant relatives (up to ninth degree) by ERSA,^19^ and then passed into PADRE^12^ to generate pedigree-aware estimates of distant relatedness.

Local IBD clustering identifies people who share an identical IBD segment spanning a specific genomic region (gene) or position (genetic variant). Since current IBD detection tools only report pairwise IBD sharing, we developed a new tool, DRIVE, to link individuals into connected graphs based on pairwise IBD sharing (Figure 1A). DRIVE is implemented in python3.6 (https://github.com/belowlab/drive). To find potential *KCNE1* D76N BioVU carriers, we used DRIVE to identify all pairwise IBD segments greater than 3 cM in length spanning *KCNE1* (Figure 1A*i*), and then conducted a three-step random walk approach, using segment length as the probability weight, to identify IBD clusters (Figure 1A*ii*). We then carried each local IBD cluster containing clinic subjects forward in analysis of potential D76N carrier clusters (Figure 1A*iii*). Finally, we used the inverse of shared IBD length to represent the local familial distance for each pair, and drew phylogenetic dendrograms with FastME 2.0 (Figure 1A*iv*).^20^

**Figure 1.**
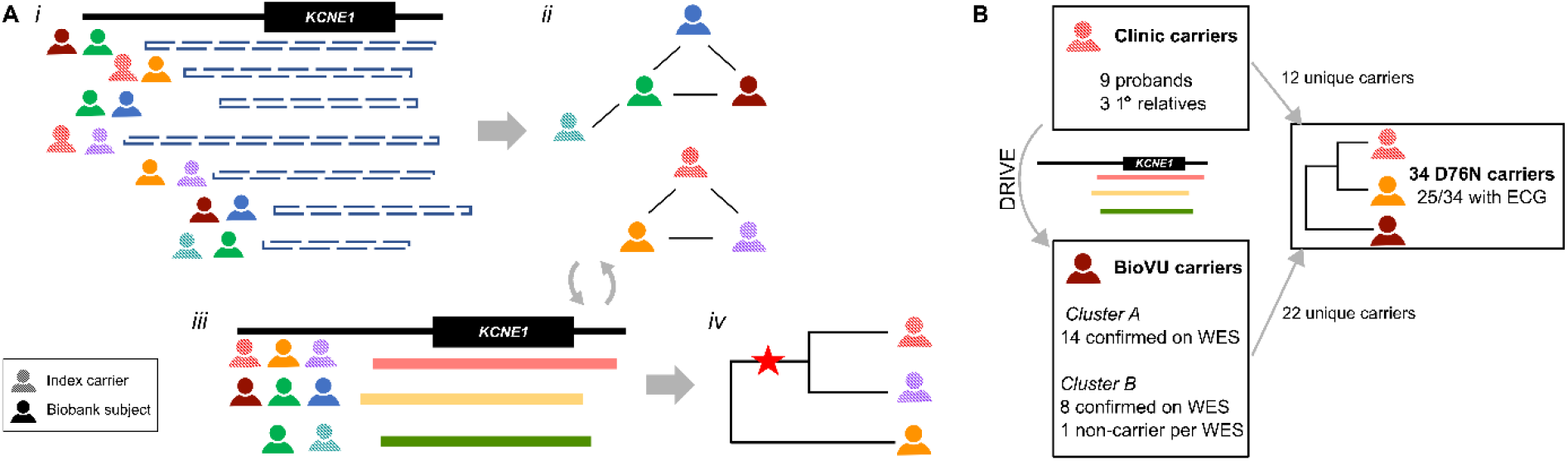
A) The DRIVE tool for local IBD clustering. This new tool identifies groups of people who share an IBD segment spanning a specific genomic region (in this study, the gene *KCNE1*). ***i***. DRIVE first selects the pairwise IBD segments spanning the target gene/variant among clinic samples and biobank subjects. ***ii***. DRIVE uses a random walk approach to cluster subjects who share the same haplotype. ***iii***. DRIVE repeats the clustering steps for large and sparse clusters. ***iv***. The inverse of the IBD segment lengths is used to represent genetic distance in a phylogenetic dendrogram. Sequence data can be integrated with the dendrogram to infer where in the family history of the genomic region the mutation event occurred (red star). **B) Study subjects**. Clinic D76N carriers comprise nine probands seen at the VUMC Genetic Arrhythmia Clinic and three related carriers identified through cascade screening. IBD-based genotype inference using the DRIVE tool was deployed in VUMC BioVU, which links the deidentified electronic health record to genomic data, to identify individuals who shared chromosomal segments at *KCNE1* with the clinic carriers. Exome sequencing confirmed D76N carrier status in 22/23 biobank individuals identified via IBD, resulting in total of 34 D76N carriers at a single center. IBD = identical by descent, WES = exome sequencing, DRIVE = Distant relatedness for Identification and Variant Evaluation. 0000-0001-6824-7155

Because the clusters showed evidence of sharing a small haplotype at *KCNE1*, suggesting co-inheritance from a common ancestor, we randomly selected two from each cluster and estimated the age of the mutation event using the recombination clock model within the Genealogical Estimation of Variant Age (GEVA) tool.^21^ The presence of D76N in each BioVU subject identified as sharing a D76N haplotype was confirmed by exome sequencing.

Genome-wide genotype imputation was performed on D76N carriers, along with 3000 BioVU controls to stabilize the imputation process, on the Michigan Imputation Server^22^ using the Haplotype Reference Consortium reference haplotypes.^23^ SNPs with low imputation quality (R^2^<0.3) were filtered. We applied the QTc PRS developed by Nauffal et al.^24^ using the *score* function in PLINK to calculate the PRS from the imputed genetic data.

ECGs obtained during routine clinical care were available for all clinic carriers, and for 13/22 biobank carriers. QTc measurements were only used if the ECG met criteria in eMethods. Arrhythmia diagnoses in each carrier’s deidentified EHR were determined using ICD-9 and ICD-10 codes and a text-search of LQTS-related terms (eMethods, eTable 2). ECG controls were BioVU subjects who clustered with the European superpopulation (European Ancestries-like, EA) in 1000 Genomes on genetic principal components analysis (n=69,819) and had an ECG read as normal (see eMethods for ECG criteria). Controls were restricted to the EA subset because all D76N carriers self-identified as White and clustered with 1000 Genomes EA population. This resulted in a control group of 3,436 genotyped individuals (2218 females,1218 males) with an ECG meeting our criteria.

Statistical tests were performed using R version 4.2.1^25^ Regression modeling used the *rms* package in R.^26^ Parametric testing was used for continuous variables if each group had >10 members and satisfied the Shapiro-Wilk test. Otherwise, non-parametric testing was used. All parametric tests were two-tailed. *P-value<*0.05 was considered statistically significant. For categorical variables, the Fisher’s exact test was used due to small sample sizes.

See eMethods for detailed methods.

## Results

### Distant relatedness in a local cluster of D76N probands

Nine D76N probands were referred to the Genetic Arrhythmia Clinic for LQTS evaluation and treatment. Cascade screening identified three additional D76N carriers among the probands’ first-degree relatives, resulting in 12 clinic subjects confirmed to carry D76N (Table 1). None of the probands were known to be related. All probands had experienced syncope, 5 had a QTc >480 ms, 5 had an implanted cardioverter-defibrillator (ICD), and 3 had documented Torsades de Pointes (TdP), the polymorphic ventricular tachycardia seen in LQTS. Among the 3 carrier family members, 1 had a QTc >480 ms, 1 had a primary prevention ICD, none had a history of syncope, and none had experienced TdP.

**Table 1.** Demographic and clinical features of D76N carriers

|  | All carriers<br>(n=34) | Clinic<br>(n=12) | BioVU<br>(n=22) | *P*-value |
| --- | --- | --- | --- | --- |
| **Demographics** |  |  |  |  |
| Age at first ECG (year) | 44.2 ± 19.7 | 42.5 ± 20.1 | 45.7 ± 19.9 | 0.69 |
| Female, n (%) | 21 (62%) | 11 (91.7%) | 10 (45.5%) | 0.011 |
| **Phenotype** |  |  |  |  |
| QTc on first ECG (msec) | 447 ± 32.9 | 463 ± 36.1 | 432 ± 21.7 | 0.019 |
| Max QTc (msec) | 463 ± 36.7 | 475 ± 33.6 | 453 ± 37.4 | 0.13 |
| Syncope, n (%) | 15 (44%) | 9 (75%) | 6 (27.2%) | 0.012 |
| Torsade or CA, n (%) | 4 (12%) | 3 (25%) | 1 (4.55%) | 0.12 |
| 1° relative with SCD or Torsade arrest | 5 (15%) | 5 (41.7%) | 0 (0%) | 0.0028 |
| **Treatment** |  |  |  |  |
| Beta blockade, n (%) | 12 (35.3%) | 9 (75%) | 3 (13.6%) | 1.7 x10<sup>-4</sup> |
| ICD, n (%) | 6 (17.6%) | 6 (50%) | 0 (0%) | 6.9 x10<sup>-4</sup> |

Data are presented as mean ± standard deviation or n (%). *P*-values compare clinic carriers to BioVU carriers, using the Fisher's exact test for categorical variables and the Mann Whitney U test for continuous variables. For age at first ECG, if no ECG was available, the current age was used. CA = cardiac arrest, SCD = sudden cardiac death, ICD= implantable cardioverter-defibrillator.

We estimated pairwise relatedness by the global proportion of IBD sharing and the distribution of shared IBD segments, using PRIMUS and ERSA software as described, enabling reconstruction of the three known nuclear pedigrees. Using PADRE, we identified previously unknown eighth to ninth degree relatedness among these pedigrees and three of the probands without close relatives (Figure 2). For reference, third cousins are eighth degree relatives. This relatedness supports the hypothesis that a local founder event underlies the comparatively high D76N frequency in Tennessee.

**Figure 2.**
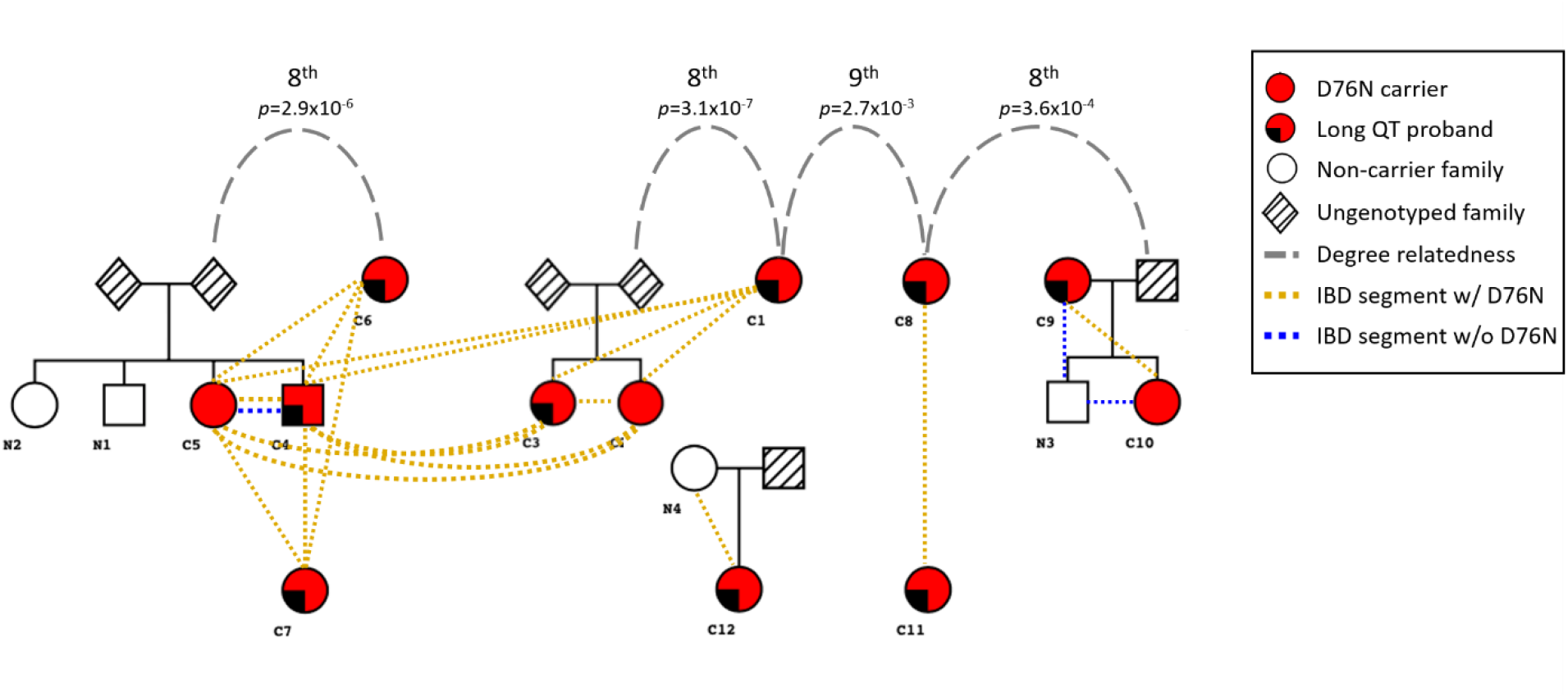
D76N probands are distantly related. Analysis of genome-wide IBD sharing in the clinic D76N carriers was used to reconstruct all known pedigrees of first-degree relatives and to identify previously unknown eighth to ninth degree relatedness among these pedigrees and three of the probands without close relatives. It is possible that more distant relatedness exists between the families that is beyond the limit of detection of existing tools (∼9^th^ degree). The colored dashed lines indicate shared IBD segments (≥ 3 cM) spanning *KCNE1*, both those harboring D76N (orange) and not (blue). IBD = identical by descent

### Using relatedness to identify D76N carriers in BioVU

We analyzed the 12 clinic carriers in conjunction with the BioVU EA genotyped population (Figure 1B). In this merged dataset, 582,671 long IBD segments (>3 cM) were detected spanning *KCNE1*. Using DRIVE, we identified 12,356 IBD clusters at the *KCNE1* locus with at least three members, including two clusters containing at least two confirmed carriers from the clinic samples. The first cluster (cluster A) included 7 clinic carriers and 14 BioVU subjects, and the second cluster (cluster B) included another 2 clinic carriers and 9 BioVU subjects. In cluster A, 82.9% of pairs shared IBD segments >3 cM (Figure 3A) spanning *KCNE1* with an average segment length of 12.3 cM. In cluster B, 88.9% of pairs shared IBD segments spanning *KCNE1* with an average length of 6.0 cM (Figure 3B). To explore more distant relatedness, we analyzed short IBD segments (>1 cM) shared across the clusters. In total, we identified 165 short IBD segments between members of cluster A and B, with an average length 1.41 cM, suggesting the two clusters are distantly related, and motivating joint analysis of variant age. Whole exome-sequencing confirmed D76N heterozygosity in 22 of the 23 BioVU subjects in the clusters. Our expanded dataset included 34 mutation carriers, 12 from clinic and 22 from BioVU. No other BioVU subjects were found to share the IBD segment (>3 cM) harboring the D76N variant. We applied a recombination clock approach, GEVA, to estimate the age of D76N. Using two BioVU carriers from each cluster and unrelated Europeans from 1000 Genomes, GEVA estimated that the mutation occurred 46 generations ago.

**Figure 3.**
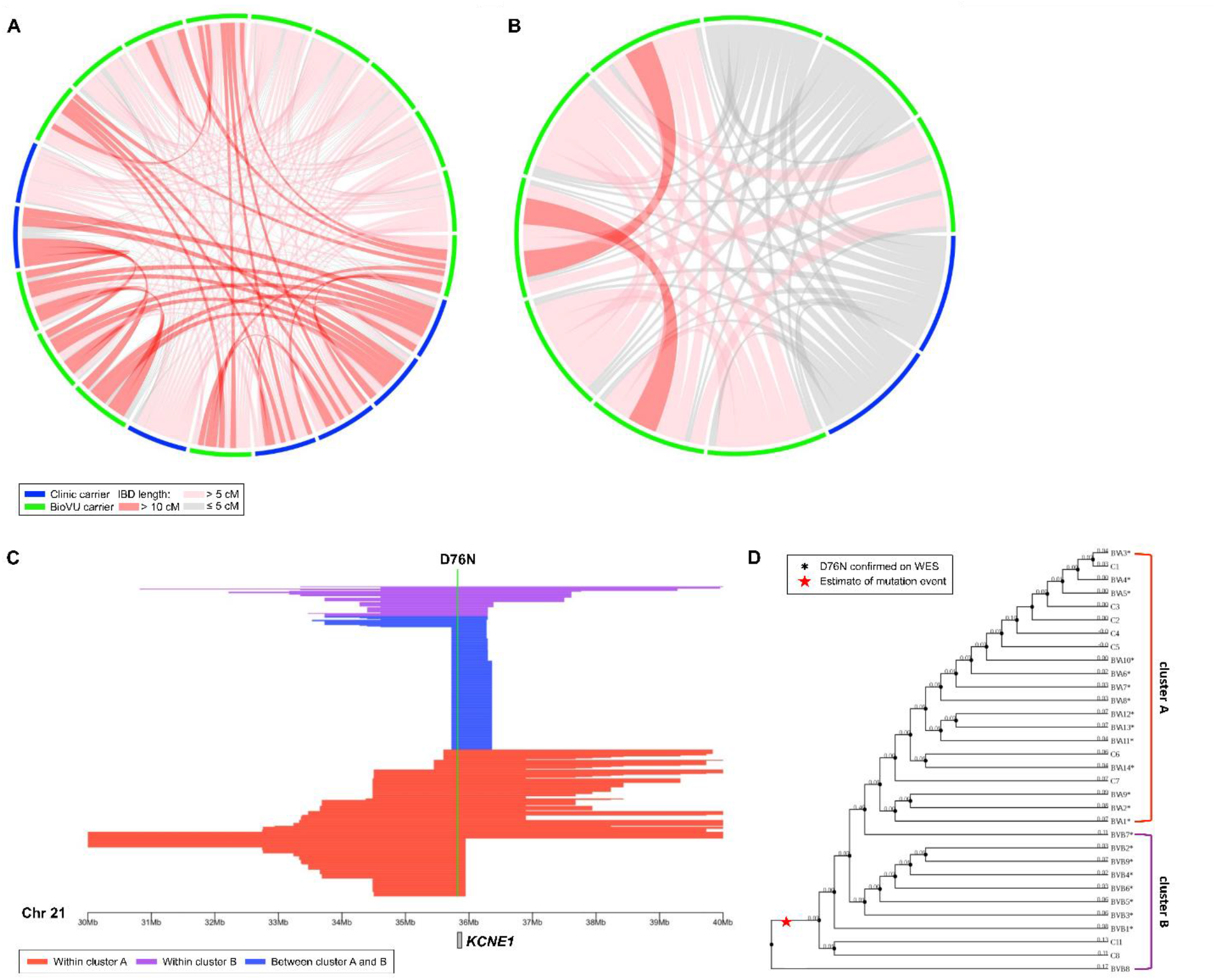
IBD clustering revealed distantly related subjects in the biobank. Local IBD sharing at *KCNE1* was analyzed between the 12 clinic carriers and the 69,879 subjects in BioVU of European descent. This identified two clusters containing known carriers, with an additional 23 subjects sharing the same chromosomal segment at *KCNE1*. Of the 23 BioVU subjects, exome sequencing confirmed D76N in 22. In the connection plots for cluster A (**A**) and cluster B (**B**), individuals are represented by the segments along the periphery. Shared chromosomal segments at *KCNE1* between individuals are indicated by the red and gray connectors. **C)** Illustration of the length and position of the shared segments spanning *KCNE1* among members of cluster A (red), members of cluster B (purple), and in members of both cluster A and B (blue). **D)** The dendrograms for cluster A and cluster B represent generational distance between subjects based on the inverse of the length of the IBD segments underlying *KCNE1* (labeled). All clinic subjects (“C” prefix) had D76N carriership determined by clinical-grade commercial testing.* indicates confirmation of D76N carriership by whole exome sequencing (WES), and the red star indicates the possible mutation event on the shared haplotype. The annotated branch length in the dendrograms for cluster A and cluster B represents the local familial distance between subjects, estimated as the inverse of the length of the IBD segments underlying *KCNE1* (numeric label on each node). IBD = identical by descent

### Cardiac events in D76N carriers

We summarized the clinical characteristics of the 34 carriers in Table 1. EHR review identified four with documented TdP. Three were subsequently seen in the Genetic Arrhythmia clinic, and are members of the clinic cohort. Of these three, one suffered an unprovoked cardiac arrest and was successfully resuscitated. The second suffered an in-hospital TdP arrest following albuterol administration. The third had self-limited TdP observed on telemetry, without arrest. TdP was documented in one BioVU carrier, attributed to sotalol use. Five clinic carriers had a documented first-degree relative with SCD or Torsades arrest. EHR review identified syncope, which is non-specific but indicates high risk in patients with LQTS,^27,28^ in 15/34 carriers. Compared to the clinic carriers, a smaller proportion of BioVU carriers were female, and fewer BioVU carriers had a history of syncope, had a first-degree relative with SCD or Torsades arrest, were on beta blockade, or had an ICD. BioVU carriers had a shorter presenting QTc compared to clinic carriers (432 msec vs. 463 msec, respectively, *p-value*=0.019).

### The QTc is prolonged in D76N carriers

Among the 25 D76N carriers with an ECG, representing both clinic and BioVU subjects, 40% of female carriers (8/20) had a QTc >480 msec, compared to 1.5% of female controls. In male carriers, 40% (2/5) had a QTc >480 msec, compared to 0.7% of male controls. The QTc, adjusted for sex, was longer in carriers (465±36 msec) compared to population controls (430±23 msec, *p-value*=4.2x10^−5^) (Figure 4A). To assess the effect of D76N in a non-referral population, we then restricted the analysis to the subset of carriers from BioVU. In this BioVU-only group, the QTc remained longer than that of population controls (456±38 msec vs. 430±23 msec, *p-value*=0.027).

**Figure 4.**
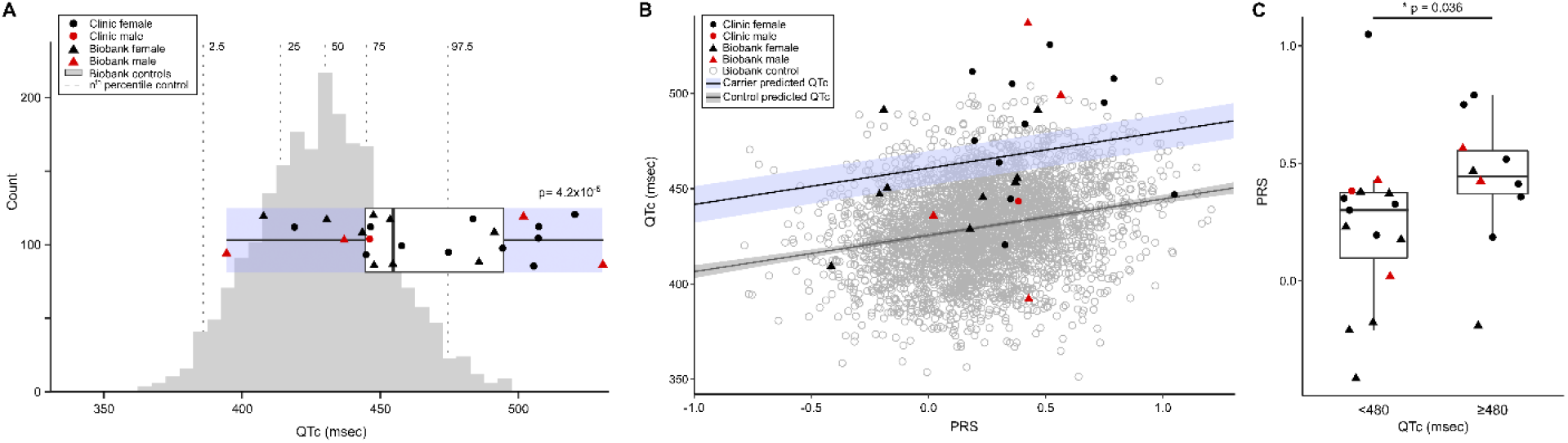
The QTc and polygenic risk in D76N carriers compared to population controls. **A)** ECGs meeting criteria detailed in eMethods were available for all clinic carriers, and for 13 of 22 biobank (BioVU) carriers. The ancestry-matched BioVU controls were selected as detailed in Methods. For both carriers and controls, if multiple ECGs were available for a subject, the maximum QTc was used. Male carrier QTcs were adjusted to female sex by adding 11.3 msec (derived from the difference between males and females in the control group when adjusted for age and PRS). The QTc was prolonged in carriers (465±6.2 msec, n=25) compared to female controls (429±22.9 msec, n= 2218; *p-value*=4.2x10^−5^). The boxplot shows the three quartiles (25%, 50%, and 75%) of the carriers. *P-*values indicate the result of the Welch unequal variances t-test between carriers and controls, with *p-value*<0.05 considered significant. **B)** Carriers have a longer QTc than controls with the same PRS. The PRS for the QTc was calculated for each carrier (clinic n=12, BioVU n=13) and for the ancestry-matched BioVU controls (n=3,436) with ECG data meeting criteria as detailed in eMethods. The predicted QTc as a function of PRS is shown for carriers (blue line) and non-carriers (gray line). **C)** Carriers with a prolonged QTc have a higher PRS than carriers with a normal QTc. The maximum QTc meeting inclusion criteria was used, and was adjusted for age and sex. The *p*-values indicates the result of Mann Whitney U test comparing carriers with maximum QTc <480, and ≥480 msec. *P-value* <0.05 was considered significant. PRS = polygenic risk score.

### Polygenic risk in D76N carriers

We assessed whether the QTc interval PRS contributed to QTc variability in D76N carriers and in controls (Figure 4B). A multivariable linear regression analysis for the QTc as a function of D76N carrier status, PRS, age, and sex was performed (eTable 3). These variables were all statistically significant in this model, which showed D76N carriers have an average 35.3 msec longer QTc compared to controls (*p-value*=1.75x10^−14^). This supports the conclusion that D76N carriers have longer QTc intervals than controls, even when polygenic risk for QT prolongation is considered. We additionally tested the interaction between PRS and D76N on QTc (eTable 3), and did not observe a significant effect beyond additive interaction (*p-value*=0.194).

We then assessed the relationship between the QTc PRS and QTc among D76N carriers. QTc intervals were adjusted for age and sex in this analysis; QTc is adjusted to female sex and age 47 (mean age of the clinic subjects), based on the regression model derived from the control population (eTable 3). The QTc PRS was different between D76N carriers with a prolonged (≥480 msec) QTc versus those without (QTc<480 msec), with a median PRS of 0.45 (interquartile range (IQR) 0.37-0.55) compared to 0.30 (IQR 0.099-0.37), respectively (*p-value*=0.036, Figure 4C). We found a trend towards higher PRS in individuals with a history of Torsades (*p-value*=0.073), and in clinic carriers compared to BioVU carriers (*p-value*=0.053) (eFigure 2A,B). The diagnosis of syncope was not associated with PRS (eFigure 2C).

## Discussion

Here we leverage patterns of sharing within the genome to make three key findings. First, we characterize both known and unknown relatedness within clinic patients identified as D76N carriers after a cardiac event (in them or a close relative) prompted referral to the Genetic Arrhythmia Clinic. The observed relatedness among carriers indicates that many carriers are descended from a common ancestor, suggesting a local founder event explains the excess of carriers of the rare D76N variant. Second, we present a novel approach in a large biobank that enabled discovery of additional D76N carriers who may be at risk of developing or have a missed diagnosis of LQT5. Third, we leverage these additional carriers from a non-referral population to evaluate the penetrance of LQT5 in D76N carriers and assess the role of QTc genetic risk factors in combination with D76N.

Both common and rare variants contribute to genetic risk in cardiovascular disease. Although the monogenic or Mendelian diseases are individually rare, in aggregate they impact 7-8% of the United States population.^29^ Since these diseases may cause high morbidity and mortality rates, early diagnosis, especially when impactful interventions are available, is especially important. Genomic sharing due to relatedness provides a powerful but underutilized approach to detect ungenotyped or poorly genotyped variation and estimate its effect. For example, Belbin *et al*.^30^ used IBD to identify multiple shared chromosomal segments associated with short stature in Puerto Ricans in an ethnically-diverse biobank, and follow-up analyses demonstrated that homozygous carriage of a rare variant in the collagen gene *COL27A1* was responsible. Although patterns of genomic sharing have been used in disease gene discovery before, the approach is not often implemented and is limited by the lack of effective tools for biobank-scale analysis.

Here we used a new approach, DRIVE, to perform biobank-scale analysis and discover 22 D76N carriers. This allowed us to determine that D76N significantly prolongs the QT interval, even in a non-referral population. In assessing pathogenicity, a commonly used criterion is allele frequency in large public datasets; however, the lack of associated phenotypic data limits the informativeness of these analyses.^31^ In the Genome Aggregation Database,^32^ the frequency of *KCNE1* D76N is very rare with a minor allele frequency of 0.00011 in Europeans, and 0.000067 overall. Thus, as with other rare variants, determination of pathogenicity and penetrance of D76N requires additional evaluation such as functional data and association with disease-related human phenotypes. In functional studies, D76N exerts profound dominant negative effects on I_Ks._ ^2^ Our human phenotypic data add further strength to the conclusion of the International Consortium that *KCNE1* variants can be monogenic causes of long QT syndrome, but penetrance is incomplete.^10^

Finally, we explored factors that modify penetrance of D76N. Polygenic variation has been shown to modulate penetrance in tier 1 monogenetic conditions^33^ as well as a rare, monogenic *SCN5A* arrhythmia syndrome.^34^ Similarly, we find an additive interaction between D76N and QT variation across the genome, as measured by a QT PRS, that suggests the risk effect of D76N is significantly modified by the rest of the genome. This work both illustrates a new paradigm for studying the effects of ungenotyped or poorly genotyped variation among cryptic relatives in regionally sampled datasets, DRIVE, and demonstrates the power of rare variant detection and evaluation in non-referral populations to improve estimates of variant effects.

In summary, our method enables identification of rare variant carriers in non-referral populations. Panel or whole genome sequencing in affected probands enables comprehensive identification of carriers of pathogenic or likely pathogenic variants in Mendelian disease genes; however, failure to detect undiagnosed carriers of rare, pathogenic variants results in missed opportunities for preventative care, and undertreatment of disease. Furthermore, failure to detect carriers in general populations with dense associated phenotypic information limits the study of rare, inherited diseases due to low sample sizes and ascertainment bias, which prevents accurate estimates of pathogenicity, penetrance, and relevant modifying effects. Most publicly available biobank data is array-based or exome-sequenced, rendering much of the functional variation unassessed. Thus, discovering subjects with shared chromosomal segments at a Mendelian disease locus known to harbor a disease-causing mutation in at least one individual holds the promise of identifying other carriers, even when their sequencing data is not available.

### Limitations

Although this study represents the largest analysis to date of the role of D76N in general populations, there are limitations to using the EHR to evaluate its effects. While none of the BioVU D76N carriers have a LQT5 diagnosis in their record, a diagnosis may have been made outside Vanderbilt. Similarly, no BioVU carriers had documentation of a first-degree family member with SCD; this could be due to insufficient family history documentation in the EHR, in contrast to the clinic carriers seen in a Genetic Arrhythmia clinic, where obtaining family history is a priority. Further, our estimation of the effects of QTc polygenic risk in carriers was limited to those with an ECG, which reduced our power and may introduce ascertainment bias. Finally, confounding by population haplotypes, genotyping error, and genotype data density limits the ability to accurately estimate the degree of relatedness using very short segments, and as a result our ability to estimate segments smaller than 3 cM, or relationships more distant than 8^th^ or 9^th^ degree.

## Conclusion

We introduce a new approach to leveraging distant relatedness to identify rare variant carriers and use this approach to identify D76N carriers who are undiagnosed for or at risk of developing LQT5. We use the set of non-referral carriers to improve estimation of D76N penetrance, detect polygenic effects modifying pathogenesis, and estimate mutation age. This demonstrates that analysis of shared chromosomal segments in large numbers of subjects with dense phenotypic data enables the discovery of mutation carriers and evaluation of disease loci.

## Data Availability

All data produced in the present study are available upon reasonable request to the authors.

## Article Information

### Affiliations

Vanderbilt University Medical Center, Nashville, TN (M.C.L, H-H.C., M.B.S, M.R.F., J.T.B., D.C.S., D.M.R., J.E.B).

University of Texas MD Anderson Cancer Center, Houston, TX (C.D.H.)

## Author contributions

Dr. Chen, Mr. Baker, Dr. Samuels, Dr. Huff, and Dr. Below developed methods.

Dr. Polikowsky participated in genetic data quality control and imputation.

Dr. Lancaster, Dr. Chen, Dr. Shoemaker, Dr. Fleming, and Dr. Roden participated in dataset acquisition.

Dr. Lancaster, Dr. Chen, Dr. Roden, and Dr. Below analyzed the data.

Dr. Lancaster, Dr. Chen, Dr. Roden, and Dr. Below wrote the manuscript.

All authors reviewed and approved the final manuscript.

## Code availability

DRIVE is available at https://github.com/belowlab/drive

