## Supplemental materials for "Detection of distant familial relatedness in biobanks for identification of undiagnosed carriers of a Mendelian disease variant: application to Long QT syndrome"

**Supplementary materials**

eMethods

eFigure 1: Familial relatedness in BioVU

eFigure 2: Comparison of the QT interval PRS

eTable 1: QT-prolonging drugs

eTable 2: Diagnostic codes used in subject phenotyping

eTable 3: Multivariable linear regression models for QTc

eReferences

**eMethods**

**Clinical samples**

Nine probands and three family members from the Genetic Arrhythmia Clinic at VUMC were clinically genotyped and found to be heterozygous carriers of the *KCNE1* D76N variant. Carrier status of 11 of these patients had been previously determined by clinical testing with commercial panels. One proband was identified incidentally in the eMERGE III (Electronic Medical Records and Genomics Phase III) sequencing study,^1^ and was subsequently referred to the Genetic Arrhythmia Clinic. All clinic carriers were of European descent.

**Biobank subjects**

BioVU is a VUMC biorepository linking deidentified electronic health records (EHRs) to over 300,000 DNA samples derived from specimens about to be discarded after clinical testing.^2^ The EHR at VUMC contributes specimen-linked de-identified demographic data, clinical notes, electronic orders, laboratory measurements (including ECG data), Current Procedures Terminology (CPT) codes, and International Classification of Diseases (ICD-9 and ICD-10) codes. Currently in BioVU, 95,124 individuals have been genotyped on the Illumina Expanded Multi-Ethnic Genomic Array (MEGA^EX^), and 54,347 of these have had at least one ECG recorded.

Controls were defined as the BioVU subjects who clustered with 1000 Genomes subjects from the European superpopulation (European Ancestries-like, EA) in genetic principal components analysis (n=69,819) and had an ECG read as normal that met the following criteria: in sinus rhythm, QRS duration 65-110 msec, heart rate 50-100 bpm, obtained in an outpatient setting with no inpatient visits immediately before or after, no QT-prolonging drugs prescribed at the time of the ECG (see Supplementary Table 1 for list of drugs^3^), no prior heart disease ICD-9 or ICD-10 codes, and potassium within the VUMC lab normal limits. Controls were restricted to the EA subset because all carriers self-described as Caucasian and clustered with 1000 Genomes subjects from the European super-population, as above. This resulted in a control group of 3,436 genotyped individuals (2218 females,1218 males) with an ECG meeting our criteria.

**MEGA^EX^ Genotyping and Quality Control**

Variants with >2% missingness and individuals with >3% missingness were excluded from further analysis. All BioVU subjects were projected against principal components from all populations in 1000 Genomes^4^ to determine genetic ancestries using PLINK.^5,6^ In total, 69,819 subjects clustered with the European superpopulation (EUR), 15,603 with the African superpopulation (AFR), 897 with the East Asian superpopulation (EAS), 414 with the South Asian superpopulation (SAS), and 2,466 with the Admixed Americans superpopulation. Within groups, additional quality control was conducted using the following thresholds for inclusion: minor allele frequency (MAF) > 0.01, variant missingness < 0.05, sample missingness <0.1, heterozygosity F<0.2, Hardy-Weinberg equilibrium p-value>1×10^-10^, removal of duplicate samples, and a requirement for genetic sex to match clinical records. After these quality assessments, 69,819 EA individuals with 718,367 variants were kept in further analysis. Although *KCNE1* D76N was genotyped by the MEGA^EX^ array, it was excluded from downstream analysis as it did not pass MAF threshold (rs74315445 MAF=0.00011 in Europeans^7^) in quality control (QC).

The 12 clinic samples were genotyped on the MEGA array to generate haplotype data using the same array and following the same protocols as those in BioVU. For these 12 subjects, written consent for genotyping was obtained under VUMC IRB approval (#9047). Because all 12 are of European ancestry and bias that can be introduced in small and highly related samples, we carried forward the set of QC-passed genetic variants in the BioVU EA dataset rather than conducting variant-based QC separately in the clinical subjects. For individual-level QC, all clinic samples’ genetic sex matched the recorded sex, heterozygosity levels were below 0.2, and all had a call rate >0.989.

**Phasing and IBD detection**

Phasing, the process of determining haplotypes (the sequence of alleles on a single chromosome) from genetic data, is a critical step for accurately identifying IBD segments. SHAPEIT4^8^ was used to establish phase in genotype data from both BioVU EA subjects and clinic samples, separately. The BioVU EA dataset is sufficiently large to conduct phasing without an external reference panel. For the 12 clinic samples, the phased BioVU EA genetic data were used as the reference panel during phasing. The BioVU EA subjects and clinic samples were merged after phasing using BCFtools.^9^ Because identification of long IBD shared segments in biobank-scale databases is a time- and computationally-intensive process, we leveraged Hap-IBD,^10^ a seed-and-extension approach, to detect IBD segments efficiently on biobank-scale data. We required a minimum shared segment length of 100 contiguous genetic markers and a minimum length of 2 centimorgan (cM; 2 cM is approximately 2 million base pairs in human) as initial seeds in Hap-IBD and carried IBD segments longer than 3 cM forward in analysis to minimize analysis of erroneous segments.

**Pedigree reconstruction and distant relatedness estimation**

Genome-wide IBD proportions were calculated using the method-of-moments estimation function in PLINK after removing ancestry-informative SNPs in PRIMUS.^11^ Non-directional networks of first- and second-degree related individuals were reconstructed into pedigrees using PRIMUS. In addition, the length and distribution of IBD segments genome-wide were used to identify more distant relatives (up to ninth degree) by ERSA,^12^ and then passed into PADRE^13^ to generate pedigree-aware estimates of distant relatedness. Relatedness estimation and pedigree reconstruction were conducted in BioVU EA subjects and clinic samples separately.

**Local IBD clustering**

The purpose of local IBD clustering is to identify groups of people who share an identical IBD segment spanning a specific genomic region (gene) or position (genetic variant). Since current IBD detection tools only report pairwise IBD sharing, we developed a new tool, DRIVE, to link individuals into connected graphs based on pairwise IBD sharing (Figure 1). To find potential *KCNE1* D76N BioVU carriers, we used DRIVE to identify all pairwise IBD segments greater than 3 cM in length spanning *KCNE1* as input (Figure 1A), and then conducted a three-step random walk approach considering the segment length as the probability weight to identify IBD clusters (Figure 1B). Random walk is an efficient approach for determining highly connected clusters within large and sparse networks, such as those derived from IBD sharing among biobank participants.^14^ Networks of close relatives would be expected to be highly connected, however spurious networks or networks connecting very short segments (due to distant relatedness) may be more sparsely connected. Therefore, for large (n > 30) and sparse (proportion of connected edges < 0.5) clusters, we ran additional random walks to split the clusters into smaller but more highly connected sub-clusters, with a maximum of five iterations of this process. We then carried each local IBD cluster containing clinic subjects forward in analysis of potential *KCNE1* D76N carrier clusters (Figure 1C). Finally, we used the inverse of shared IBD length to represent the local familial distance based on haplotypes shared at the *KCNE1* locus for each pair and drew phylogenetic dendrograms with FastME 2.0 (Figure 1D).^15^

**Whole exome-sequencing validation**

The presence of *KCNE1* D76N in each BioVU subject identified as sharing a D76N haplotype was confirmed by exome sequencing on an Illumina NextSeq 500 with 150 bp paired-end reads following a standard Illumina protocol by the sequencing core at VANTAGE (Vanderbilt Technologies for Advanced Genomics). Sequencing quality control was conducted by fastp to filter out short, uncertain, and low-quality reads.^16^ Reads that passed QC were aligned to the hg38 human genome reference by BWA2,^17^ and the exome-wide variants were called by GATK following standard best practices.^18^

**Mutation age estimation**

From the newly identified BioVU D76N carriers, because the clusters showed evidence of sharing a small haplotype at *KCNE1*, suggesting co-inheritance from a very common ancestor, we randomly selected two from each cluster (Figure 5B) and estimated the age of the mutation event using the recombination clock model within the Genealogical Estimation of Variant Age (GEVA) tool.^19^ The length of shared segments spanning the target variant were used to estimate the time to the most recent common ancestor with the European ancestry 1000 Genomes data as reference.^4^ GEVA then estimates the age of the mutation event by comparing the estimated time to the most recent common ancestor among the pair of subjects that carry the target variant relative to pairs in which only one subject carries the target variant and the other one does not.

**Genome imputation and polygenic risk score calculation**

Genome-wide genotype imputation was performed on D76N carriers along with 3000 BioVU controls to stabilize the imputation process. The imputation controls were randomly selected from the subset of BioVU EA subjects with MEGA genotypes, described above. The subset of single nucleotide polymorphisms (SNPs) passing quality control that overlapped between D76N carriers and the imputation controls were used in imputation on the Michigan Imputation Server^20^ using the European ancestry Haplotype Reference Consortium version r1.1 2016 reference haplotypes.^21^ SNPs with low imputation quality (R^2^<0.3) were filtered before further analysis.

We then used the PRS for QTc developed by Nauffal et al.^22^ This PRS comprises 1,110,494 SNPs, and there were 1,110,297 overlapping SNPs between the PRS and our imputed genomes. Using the *score* function in PLINK, we calculated the PRS from the imputed genetic data of each carrier and control subject.

**Electronic health records review**

ECGs obtained as part of routine clinical care were available for all clinic carriers, and for 13 of 22 biobank carriers. Correction for heart rate was performed using the Bazett formula. For carriers, QTc measurements were only used if the ECG was not obtained while inpatient, no QT-prolonging drugs were prescribed at the time of the ECG (Supplementary Table 1), was in sinus rhythm or atrially paced, had a QRS duration <110 ms, and had a heart rate 50-100 bpm. The reference QTc distribution for males and females was derived from the control group described above.

Arrhythmia diagnoses in BioVU carriers were determined from ICD-9 and ICD-10 codes in their deidentified EHR; a full list of codes used is presented in the Supplemental Table 2. Additionally, a text-search of each subject’s EHR was performed for the terms “cardiac arrest,” “long QT,” “LQT,” “PMVT,” “polymorphic ventricular tachycardia,” “SCD,” “seizure,” “sudden cardiac death,” “syncope,” “Torsade(s),” “TdP,” “ventricular fibrillation,” “V fib,” and “VF;” any matches were manually reviewed to confirm the diagnosis.

**Statistical analysis**

All statistical tests were performed using R version 4.2.1^23^ and the *dplyr* package.^24^ Regression modeling was done with the *rms* package in R.^25^ For continuous variables, parametric testing was used if each group being compared had >10 members and satisfied the Shapiro-Wilk test of normality. Otherwise, non-parametric testing was used. All parametric tests were two-tailed. *P-value<*0.05 was considered statistically significant. For categorical variables, the Fisher’s exact test was used due to small sample sizes. The *ggplot2* package was used in figure preparation.^26^

**eFigure 1**

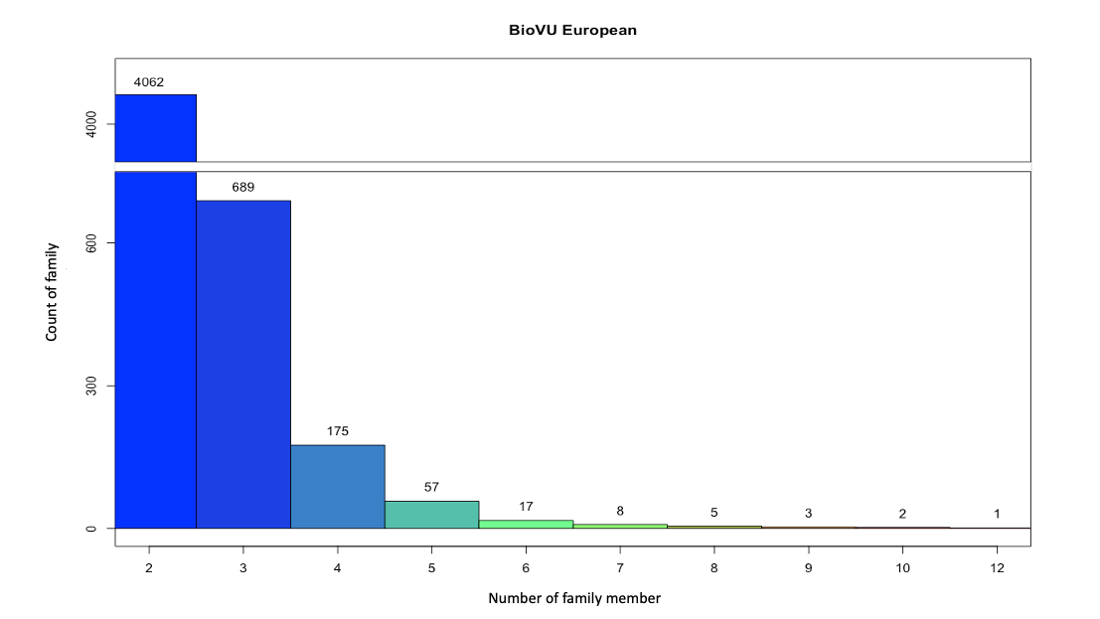

**Familial relatedness in BioVU.** Histogram showing the count of families in BioVU connected by first- and second-degree relationships, ordered by size. That is, there are 4062 families in BioVU connected by first- and second-degree relationships that contain two people, and there is one family containing 12 people.

**eFigure 2**

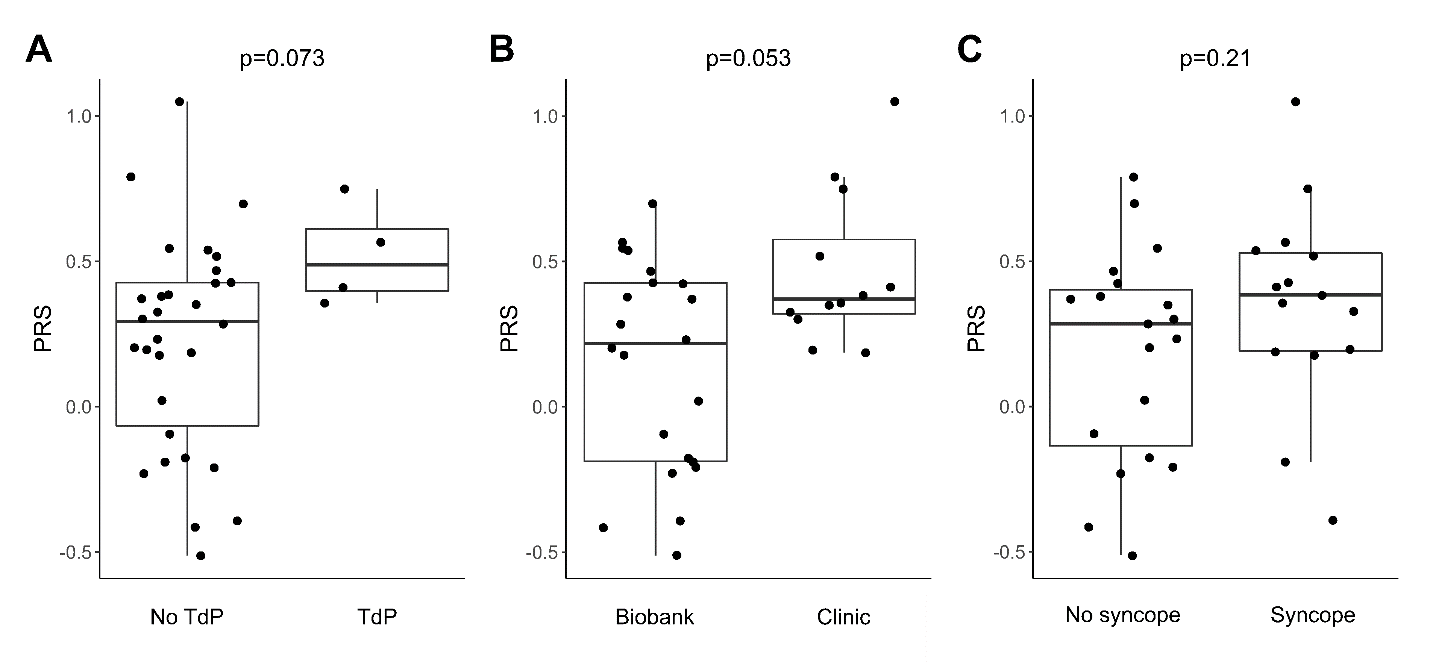

**Comparison of the QT interval PRS in A.** D76N carriers with a history of Torsades vs. those without Torsades, **B.** D76N carriers from clinic (the referral group) vs. carriers from the biobank (the non-referral group), and **C.** D76N carriers with a history of syncope vs. those without syncope. *P*-values indicate the results of Wilcoxon rank-sum test. *P* < 0.05 was considered significant. PRS = polygenic risk score, TdP = Torsades de Pointes.

**eTable 1. QT-prolonging drugs**

| **Drug name** | **Drug name** |
| --- | --- |
| Aclarubicin | Ibogaine |
| Amiodarone | Ibutilide |
| Anagrelide | Levofloxacin |
| Arsenic trioxide | Levomepromazine, methotrimeprazine |
| Astemizole | Levomethadyl acetate |
| Azithromycin | Levosulpiride |
| Bepridil | Meglumine antimoniate |
| Cesium Chloride | Mesoridazine |
| Chloroquine | Methadone |
| Chlorpromazine | Mobocertinib |
| Chlorprothixene | Moxifloxacin |
| Cilostazol | Nifekalant |
| Ciprofloxacin | Ondansetron |
| Cisapride | Oxaliplatin |
| Citalopram | Papaverine |
| Clarithromycin | Pentamidine |
| Cocaine | Pimozide |
| Disopyramide | Probucol |
| Dofetilide | Procainamide |
| Domperidone | Propofol |
| Donepezil | Quinidine |
| Dronedarone | Roxithromycin |
| Droperidol | Sertindole |
| Erythromycin | Sevoflurane |
| Escitalopram | Sotalol |
| Flecainide | Sparfloxacin |
| Fluconazole | Sulpiride |
| Gatifloxacin | Sultopride |
| Grepafloxacin | Terfenadine |
| Halofantrine | Terlipressin |
| Haloperidol | Terodiline |
| Hydroquinidine, dihydroquinidine | Thioridazine |
| Hydroxychloroquine | Vandetanib |

QT-prolonging drugs in the “Known Risk of Torsades” category in the CredibleMeds database.^3^

**eTable 2. Diagnostic codes used in subject phenotyping**

| **ICD-9 code** | **Description** |
| --- | --- |
| 426.82 | Long QT syndrome |
| 427.42 | Ventricular flutter |
| 427.1 | Paroxysmal ventricular tachycardia |
| 427.41 | Ventricular Fibrillation |
| 427.5 | Cardiac arrest |
| 798.1 | Instantaneous death |
| 798.2 | Death occurring in less than 24 hours from onset of symptoms, not otherwise explained |
| 798.9 | Unattended death |
| V12.53 | Personal history of sudden cardiac arrest |
| 780.2 | Syncope and collapse |
| **ICD-10 code** | **Description** |
| I45.81 | Long QT syndrome |
| I47.2 | Ventricular tachycardia |
| I49.0 | Ventricular fibrillation and flutter |
| I49.01 | Ventricular fibrillation |
| I49.02 | Ventricular flutter |
| I46.9 | Cardiac arrest |
| P29.81 | Cardiac arrest (newborn) |
| R99 | Death cause unknown |
| Z86.74 | Cardiac arrest (death) successfully resuscitated, personal history |
| R55.9 | Syncope and collapse |

**eTable 3**. **Multivariable linear regression models for QTc**

|  | Beta | SE | P-value |
| --- | --- | --- | --- |
| **Model 1** | | | |
| Intercept | 417.99 | 1.38 | <2.2x10^-16^ |
| D76N | 35.26 | 4.58 | 1.75x10^-14^ |
| PRS | 19.07 | 1.34 | <2.2x10^-16^ |
| Age | 0.14 | 0.02 | 5.43x10^-9^ |
| Sex | -11.42 | 0.81 | <2.2x10^-16^ |
| **Model 2** | | | |
| Intercept | 418.06 | 1.38 | <2.2x10^-16^ |
| D76N | 29.57 | 6.33 | 3.14x10^-6^ |
| PRS | 18.90 | 1.35 | <2.2x10^-16^ |
| Age | 0.14 | 0.02 | 6.18x10^-9^ |
| Sex | -11.42 | 0.81 | <2.2x10^-16^ |
| D76N*PRS | 18.52 | 14.26 | 0.194 |
